# Genetic architecture of four smoking behaviors using partitioned *h*^*2*^_*SNP*_

**DOI:** 10.1101/2020.06.17.20134080

**Authors:** Luke M. Evans, Seonkyeong Jang, Marissa A. Ehringer, Jacqueline M. Otto, Scott I. Vrieze, Matthew C. Keller

**Author notes:** Address correspondence to: Luke M. Evans PhD, Institute for Behavioral Genetics, University of Colorado Boulder, 1480 30^th^ St., Boulder, CO 80303. **The authors declare no conflicts of interest.**.

## Abstract

**Background and Aims:** Smoking is a leading cause of premature death. Although genome-wide association studies have identified many loci that influence smoking behaviors, much of the genetic variance in these traits remains unexplained. We sought to characterize the genetic architecture of four smoking behaviors through SNP-based heritability (*h*^*2*^_*SNP*_) analyses.

**Design:** We applied recently-developed partitioned *h*^*2*^_*SNP*_ approaches to smoking behavior traits assessed in the UK Biobank.

**Setting:** UK Biobank.

**Participants:** UK Biobank participants of European ancestry. The number of participants varied depending on the trait, from 54,792 to 323,068.

**Measurements:** Smoking initiation, age of initiation, cigarettes per day (CPD; count, log-transformed, binned, and dichotomized into heavy versus light), and smoking cessation. Imputed genome-wide SNPs.

**Findings:** We estimated *h*^*2*^_*SNP*_(SE)=0.18(0.01) for smoking initiation and 0.12(0.02) for smoking cessation, which were more than twice the previously reported estimates. Estimated age of initiation *h*^*2*^_*SNP*_=0.05(0.01) and binned CPD *h*^*2*^_*SNP*_=0.1(0.01) were similar to previous reports. These estimates remained substantially below published twin-based *h*^*2*^ of roughly 50%. CPD encoding strongly influenced estimates, with dichotomized CPD *h*^*2*^_*SNP*_=0.28. We found significant contributions of low-frequency variants and variants in low linkage-disequilibrium (LD) with surrounding genomic regions. Functional annotations related to LD, allele frequency, sequence conservation, and selective constraint also contributed significantly to the partitioned heritability. We found no evidence of dominance genetic variance for any trait.

**Conclusion:** *h*^*2*^_*SNP*_ of these four specific smoking behaviors is modest overall. The patterns of partitioned *h*^*2*^_*SNP*_ for these highly polygenic traits is consistent with negative selection. We found a predominant contribution of common variants, and our results suggest a role of low-frequency or rare variants, poorly tagged by surrounding regions. Deep sequencing of large samples and/or improved imputation will be required to fully assess the role of rare variants.

## Introduction

Cigarette smoking is a leading cause of premature death worldwide(1), and many smokers struggle to quit, despite interest and numerous attempts(2). Smoking prevalence has decreased in recent decades due to public health efforts(3); however, rates of alternative forms of nicotine use (e.g., vaping) have grown rapidly during this time(4), demonstrating a pressing need to characterize the underlying biology of nicotine use and smoking to reduce subsequent premature death.

A key aspect of that underlying biology is the genetic architecture(5) of smoking behaviors, including the relative contribution of rare vs. common variants, functional annotation of associated loci, and characterization of the neutral and selective forces shaping that architecture. Numerous(6-10) twin, adoption, and family studies have demonstrated that up to 50% of the variance in nicotine dependence and individual smoking behaviors, such as quantity, is attributable to genetic influences. Recent genome-wide association studies (GWAS) have improved our understanding of this genetic basis by identifying over two hundred conditionally independent loci associated with these traits to date(11-14). This genetic signal is enriched in loci that influence the epigenome and within specific brain regions such as the hippocampus, providing a more nuanced interpretation of specific class(es) of variants, candidate brain regions, and potential causal mechanisms that influence smoking(11). Together, this body of work strongly indicates a highly polygenic architecture to smoking behaviors. Nonetheless, significantly associated loci collectively explain only a small proportion of the family-based genetic variance, leaving many additional loci undiscovered and the majority of the genetic variance unexplained.

While additional common variants of very small effect are likely to be identified as sample sizes grow, some of the unexplained variability undoubtedly arises from uncommon and rare variants (MAF<0.01), though their relative contribution is uncertain. The most recent large GWASs(11, 15) of smoking behaviors and nicotine dependence, using LD score regression (LDSC), estimate the SNP-based heritability (*h*^*2*^_*SNP*_) due to common variants as 0.05-0.09 across traits(16). A related exome sequencing study(17) estimated that rare coding variants explained approximately 1-2% of the phenotypic variance. However, given that the majority of identified associations are intergenic(11), exome-based studies are unlikely to identify most rare variants influencing these behaviors. Thus, the rare-variant contribution to smoking behaviors may yet be substantial when assessed with methods that can account for the aggregated influence of common and rare variation.

Additionally, the contribution of non-additive genetic variance to these smoking behaviors is poorly understood. Twin-based studies have typically evaluated ACE models(9), which estimate additive genetic (A), common environment (C) and unique environment (E) variances using twin correlations, implicitly assuming zero dominance genetic variance. Extended twin kinship models can estimate dominance genetic variance and shared environmental effects simultaneously, and the only such model to evaluate smoking initiation found no evidence of dominance genetic variance(18). Although several *h*^*2*^_*SNP*_ estimates of smoking behaviors have been to published, to our knowledge only one estimate of SNP-based dominance genetic variance (*δ*^*2*^_*SNP*_) has been reported, which found *δ*^*2*^_*SNP*_ of smoker status indistinguishable from zero(19). Furthermore, the allele frequency spectrum and contribution of functional annotations related to LD, allele frequency, recombination and related genomic features for smoking behaviors has not been fully explored. One study applied partitioned *h*^*2*^_*SNP*_ approaches to evaluate tissue-specific effects, with results indicating that genes expressed in the cerebellum are enriched in their contribution to nicotine dependence(15). The only published work has examined a single trait, smoking status, finding contributions of low-LD and -MAF variants consistent with negative or purifying selection(20, 21). Whether these same patterns exist for other smoking behaviors, such as quantity of use or cessation, is unknown.

A comprehensive evaluation of the frequency spectrum, the influence of dominance genetic variance, and the contributions of functional annotations is needed to provide a more complete picture of the genetic architecture underlying complex smoking behaviors. Here, we use recently developed methods(19, 21-26) to evaluate these heritable contributions and characterize the genetic architecture of four smoking behaviors: smoking initiation (whether an individual has ever been a regular smoker), age of initiation of regular smoking, cigarettes per day (evaluated with different data encodings), and smoking cessation.

## Methods

### Phenotype and Genetic Datasets

Using the UK Biobank(27) full release, we assessed the same four smoking phenotypes as the GSCAN project(11), defined identically (final sample sizes after quality control; see below): 1) smoking initiation (N=323,068), defined as whether an individual had ever in their lifetime been a regular smoker by having smoked over 100 cigarettes over one’s lifetime; 2) age of smoking initiation (N=122,200), defined as the age at which an individual began smoking regularly (UK Biobank data fields 3426 and 2867); 3) cigarettes per day (CPD; N=116,258), defined as a 5-bin variable based on responses for the number of cigarettes smoked per day (fields 2887, 3456, and 6183); and 4) smoking cessation (N=160,390), defined as individuals who were not current smokers but had been regular smokers at one point (fields 1239 and 1249). The latter three phenotypes required an individual to be a current or former regular smoker. GREML variance estimation (see below) was limited by available RAM (1 Tb) on a single compute node; therefore, we analyzed the smoking initiation and smoking cessation data using three and two separate, equally sized subsamples, respectively, and meta-analyzed the results using inverse-variance weighting. Age of initiation and CPD were each analyzed in a single analysis. In addition to the binned CPD metric used in recent genetic association meta-analyses(11), we examined the influence of CPD scale on *h*^*2*^_*SNP*_ estimates, which we previously found to influence association effect sizes(28). We evaluated raw CPD count, log-transformed CPD, √CPD, CPD^(2/3)^, and dichotomized CPD (heavy vs. light) using four different sets of CPD cutoffs for heavy and light smoker definitions (we applied the following Heavy (H) and Light (L) cutoffs of CPD: a) H: >20, L: <=10; b) H: >30, L: <=10; c) H: >40, L: <=5; d) Median CPD of 20 (H: >20, L: <=20); Figure S1). Final sample sizes for the different CPD encodings are presented in the Supplemental Information. These phenotypes encompass key aspects of nicotine dependence(29).

The UK Biobank release included ∼97M imputed variants using both the Haplotype Reference Consortium (HRC) and 1000 Genomes+UK10K reference panels(27). We removed individuals with mismatched self-reported and genetic sex, |F_het_|≥0.2, and/or no phenotypic information. We restricted our analyses to biallelic SNPs with minor allele frequency (MAF)≥0.0001, imputation INFO score≥0.3, Hardy-Weinberg equilibrium test (HWE) p-value≥10^−10^, and variant missingness≤0.02 using plink1.9(30), yielding 22,982,114 SNPs. The choice of INFO score threshold was based on previous results demonstrating that variants with relatively poor imputation still contribute to *h*^*2*^_*SNP*_ estimates(23), although *h*^*2*^_*SNP*_ for a given partition will be underestimated to the degree that SNPs in that bin have average INFO<1 (22). We identified individuals of European ancestry using principal components analysis using flashpca (31) from a set of MAF- and LD-pruned array markers (plink2 command: --maf 0.05 --indep-pairwise 50 5 0.2), retaining those whose scores on the first four PCs fell within the range of the UK Biobank-identified individuals of European ancestry (UK Biobank data field ID 22006). We identified unrelated individuals using GCTAv1.91.3 (32) with an initial relatedness cutoff of < 0.05. After observing differences between REML- and Haseman-Elston-based variance estimators (see below), we applied relatedness thresholds of 0.02, 0.03, 0.04 & 0.05 to assess the potential for environmental effects confounding rare variation. Because sample size varied for each of the four phenotypes, we applied these relatedness thresholds for each phenotype separately. All sample sizes are presented in Tables S1-S3.

### Variance Estimation

We estimated genetic variance in unrelated individuals using a set of genetic relatedness matrices (GRMs) partitioned by MAF- and individual marker LD-stratified bins (LDMS-I), which provides the most robust estimates of genetic variance across the allelic frequency spectrum in imputed data(22) and can be used in a GREML (GCTA(32)) or moment-matching framework such as phenotype correlation-genotype correlation (PCGC) regression(33, 34). These analyses were not pre-registered, and are therefore exploratory. We used both GCTA and PCGC (for binary traits) to estimate variances accounted for by GRMs (described next), and included the following as fixed effect covariates: sex (UK Biobank field ID 31), age (21003), age^2^, Townsend deprivation index (189), educational attainment (6138), genotyping batch (22000), scores of the first 10 worldwide principal components (22009), and scores of the first 10 principal components of the retained individuals of European ancestry estimated as described above.

We estimated *h*^*2*^_*SNP*_ using six LDMS-I-partitioned GRMs. We calculated LD scores for all imputed markers (GCTA: --ld-score-region 200). We stratified markers into four MAF intervals ([0.0001, 0.001), [0.001, 0.01), [0.01, 0.05), ≥0.05). For the two more common MAF bins, we further stratified SNPs into low and high individual SNP LD score bins based on median LD score within MAF bins. We did not LD stratify the rarest two MAF bins because there is low variation in LD for low MAF SNPs (most SNPs have low LD), because of limited power to differentiate across LD bins of SNP s of low MAF, and because inclusion of more GRMs required more memory than available. Because of incomplete data across all four phenotypes, we estimated all GRMs for each set of unrelated individuals for each phenotype separately.

To estimate dominance genetic variance, *δ*^*2*^_*SNP*_, we included a dominance genetic relatedness matrix(19) for each dataset (GCTA: --make-bin-d) using all markers with MAF>0.01. We did not partition the dominance matrix by MAF or LD because of practical limitations, noted above.

For binary traits (age of initiation, smoking cessation, and heavy/light CPD), we converted observed scale *h*^*2*^_*SNP*_ estimates to the liability scale using within-sample trait prevalence and the conversion of Lee et al.(35).

Finally, we evaluated the influence of the relatedness threshold used, i.e., potential environmental confounding and cryptic relatedness, by using progressively lower relatedness thresholds (0.02, 0.03, 0.04 and 0.05), then estimating *h*^*2*^_*SNP*_ as above. Resulting sample sizes across thresholds are presented in Tables S1-S3.

### Functional Annotation and Tissue-Specific Expression Heritability Enrichment

We used LD Score Regression to estimate partitioned *h*^*2*^_*SNP*_ for functional annotations (25). We applied the baseline+LD model (21) to assess functional annotations such as LD, allele frequency and age, recombination rate, and related annotations, and the possible role of purifying selection. We applied a Bonferroni cutoff either within traits (*p*<0.00052, as suggestive) or across all traits (*p*<0.00013) to identify significant LDSC regression coefficients.

## Results

Using GREML-LDMS-I with unrelated individual, we estimated smoking initiation *h*^*2*^_*SNP*_(SE)=0.176(0.007), smoking cessation *h*^*2*^_*SNP*_=0.119(0.018), cigarettes per day *h*^*2*^_*SNP*_=0.098(0.011), and age of initiation *h*^*2*^_*SNP*_=0.055(0.011) (Figure 1, Table S1). MAF- and LD-partitioned heritability estimates differed across traits. Common variants (MAF>0.05) contributed substantially to all traits, particularly common variants with relatively low LD (Figure 1). Uncommon variants (MAF 0.01-0.05) with low LD, but not high LD, contributed to all traits. Alternatively, uncommon (MAF<0.01) variants contributed significantly only to smoking initiation and age of initiation, and rare (MAF<0.001) variants did not contribute significantly to any trait.

**Figure 1.**
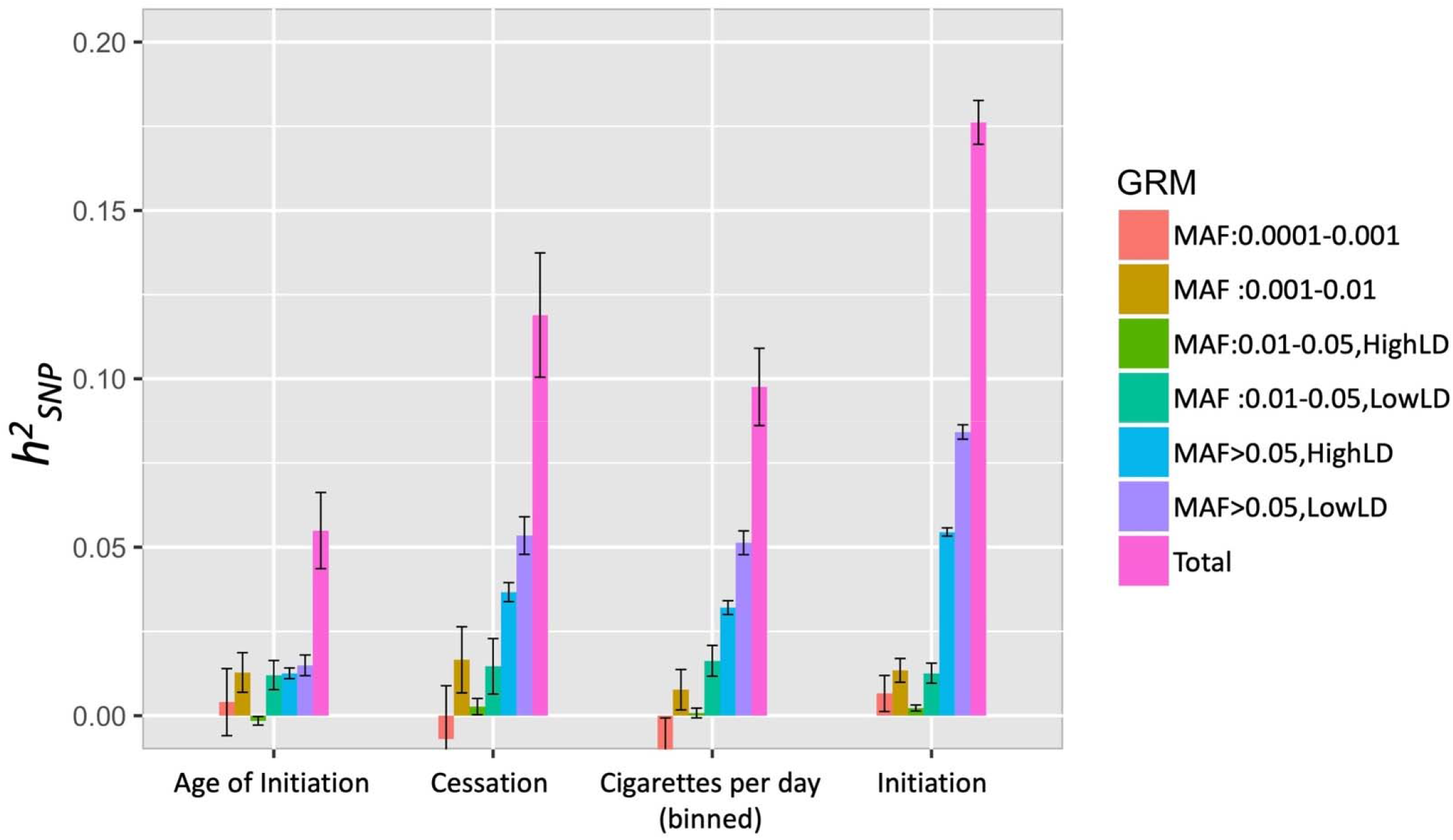
*h*^*2*^_*SNP*_ estimates (+/- standard error) across four smoking behaviors, partitioned using GREML-LDMS-I. Note that twin-based estimates are roughly 50% across these smoking traits.

Notably, we estimated significantly different (non-overlapping 95% CI) total and binned *h*^*2*^_*SNP*_ for different CPD encodings. Total *h*^*2*^_*SNP*_ ranged from 0.092(0.011) for the raw CPD count to 0.289(0.038) when CPD was dichotomized into heavy(CPD>20)/light(CPD<=10) smokers (Figures 2 & S2-S3, Tables S1-3). All dichotomized CPD total *h*^*2*^_*SNP*_ estimates (except using the median) were >0.2. We found differences in partitioned estimates, where common variants (MAF>0.05) contributed to substantially higher *h*^*2*^_*SNP*_ of heavy/light CPD than the other CPD encodings. Rarer (MAF 0.001-0.01) variant contribution was also higher, though the smaller sample size of the dichotomized data led to larger standard errors.

**Figure 2.**
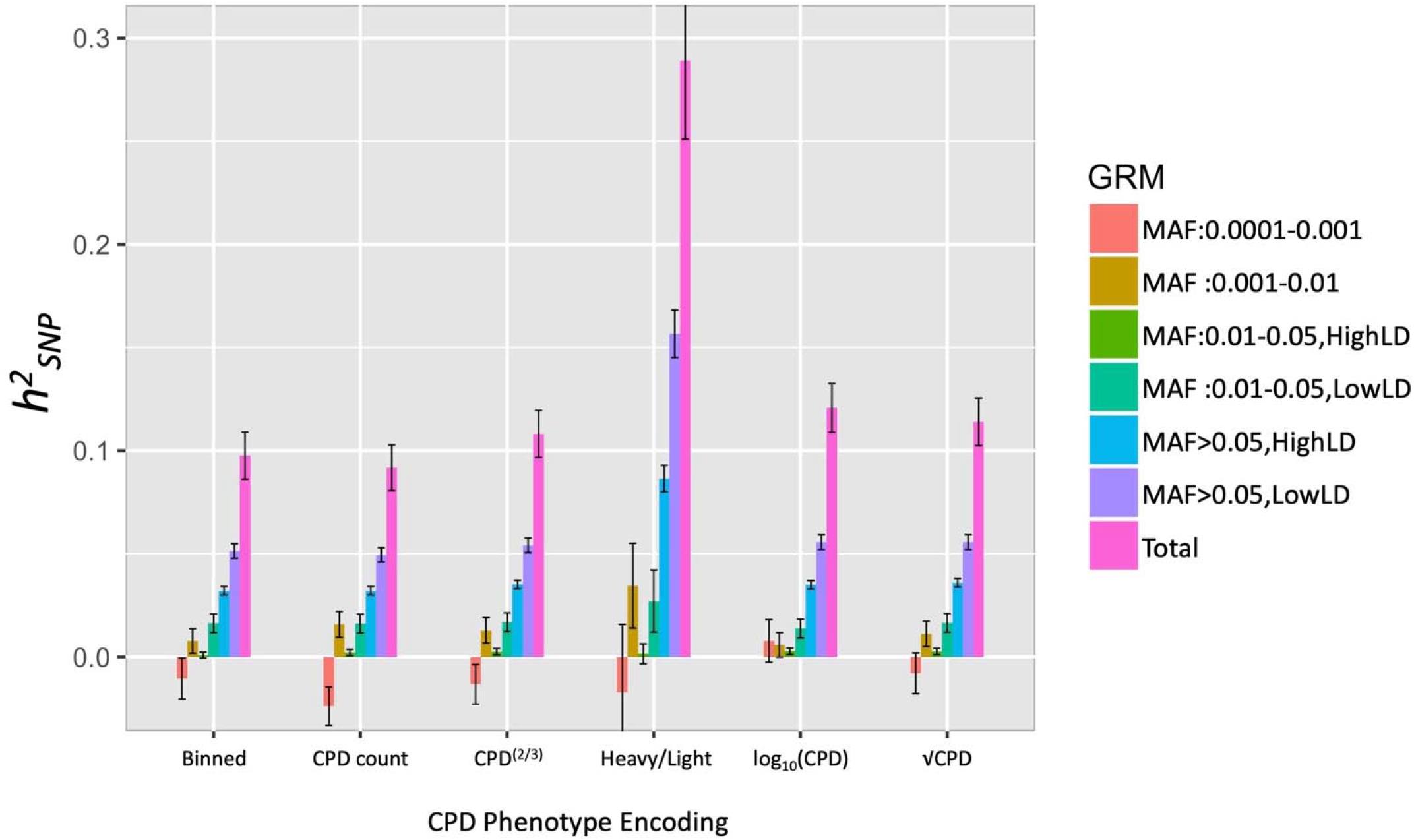
*h*^*2*^_*SNP*_ estimates (+/- standard error) of CPD using different phenotype encodings, partitioned using GREML-LDMS-I. Heavy vs. light is dichotomized with Light: CPD<=10 and Heavy: CPD>20; estimated *h*^*2*^_*SNP*_ shown on the liability scale using a prevalence of 0.42.

We estimated the contribution of dominance variance. For all traits, the 95% CI of *δ*^*2*^_*SNP*_ estimates overlapped zero (Table S4).

The relatedness threshold strongly influenced estimated *h*^*2*^_*SNP*_ when using PCGC, but not when using GREML (Tables S1-3, Figs. S2-S6). Specifically, the PCGC estimates were considerably higher than GREML estimates when applying a relatedness<0.05 cutoff with smoking initiation and smoking cessation, but dropped and had overlapping 95% CIs at lower relatedness thresholds. The higher estimates when using PCGC with relatedness<0.05 were driven by a much greater contribution of rare variant *h*^*2*^_*SNP*_ (MAF<0.0001; Figure S5-S6).

We applied partitioned LDSC to assess contribution of functional annotations and the role of LD and selective constraint in smoking behaviors. Across smoking behaviors, we found that SNPs that were highly conserved, that had lower MAF-adjusted LD or lower MAF quantiles (MAF>0.001 in Liu et al.(11)), and that were in areas of high CpG content and low recombination rate contributed significantly to heritable genetic variation (Figures 3 & S7, Table S5).

**Figure 3.**
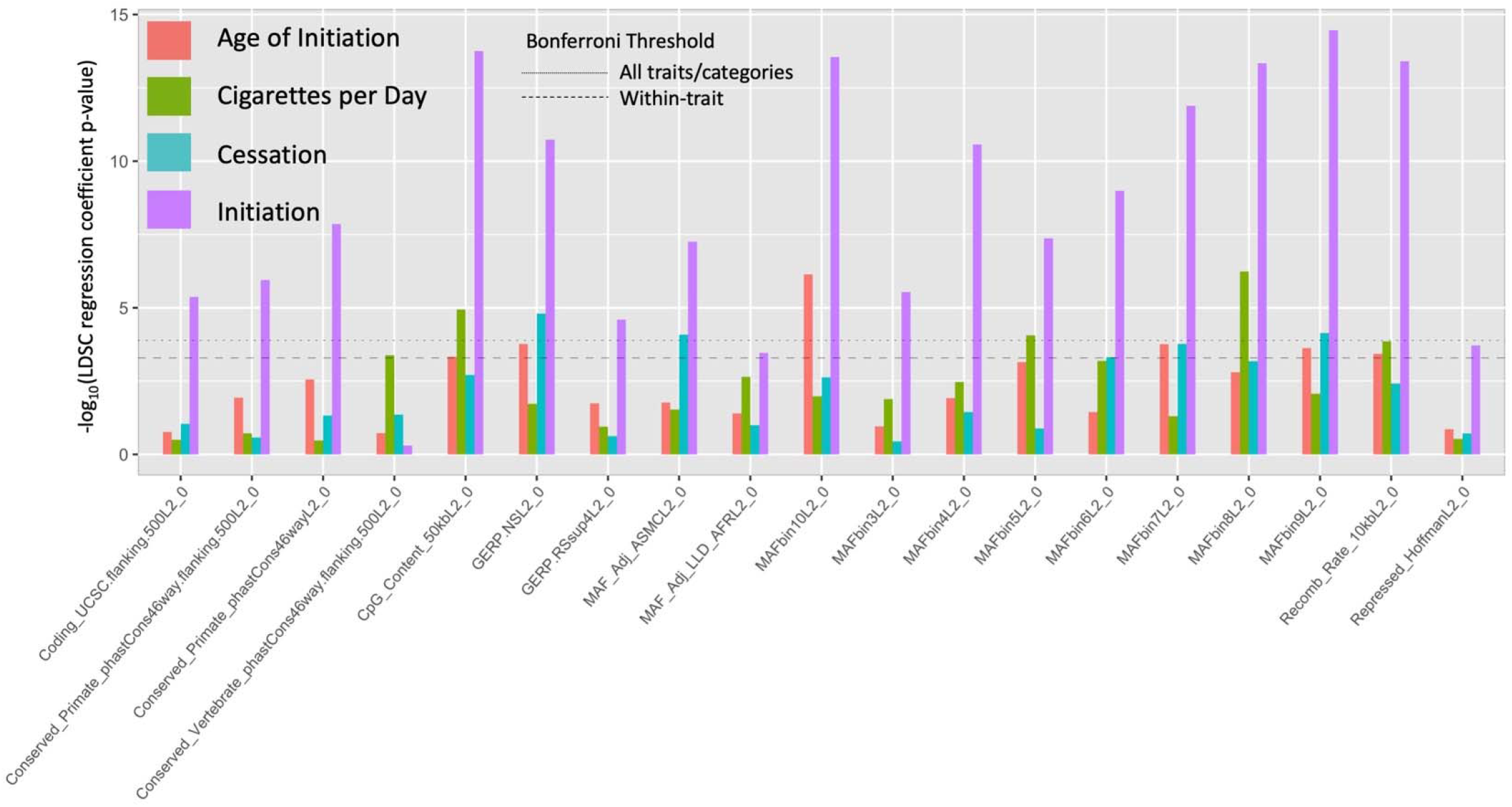
Partitioned LDSC regression coefficient p-values for all annotations with at least one significant coefficient across all traits. See Supplemental Figure S5 & Table S3 for all annotations.

## Discussion

We estimated *h*^*2*^_*SNP*_ and *δ*^*2*^_*SNP*_ across four key smoking behaviors, and partitioned variance among rare vs. common variants and functional annotations. Our *h*^*2*^_*SNP*_ estimates are more than double the previously reported(11) LDSC-based and single-component GREML-based estimates for smoking initiation (0.18 vs. 0.08 and 0.12) and smoking cessation (0.12 vs. 0.05 and 0.06), but are nearly identical for binned CPD (0.1). Our estimate of age of smoking initiation *h*^*2*^_*SNP*_=0.05 is nearly identical to the LDSC-based estimate, but is much lower than the previous single-component GREML estimate of 0.11. The difference in age of initiation *h*^*2*^_*SNP*_ may be due to including all variants in a single GRM when the causal variants are relatively common(22). Partitioned estimates of common, well-tagged variants are similar to the LDSC-based estimates(11) across all four traits, consistent with expectations, as LDSC estimates variance due to common, well-tagged variants(16, 22). The higher *h*^*2*^_*SNP*_ estimates for smoking initiation and cessation results from larger contributions of low-LD and low-frequency variants (MAF<0.01), suggesting that for these traits, a non-trivial portion of the genetic variance is due to rarer variants and those that are poorly tagged by surrounding SNPs. This contribution is likely underestimated in the current study, because even with HRC-imputed data, these sites are typically poorly imputed, which leads to a downward bias in *h*^*2*^_*SNP*_ estimates(22, 23).

Alternative CPD encodings led to different estimates, wherein total *h*^*2*^_*SNP*_ for dichotomized heavy/light smoker status was over twice that of other encodings. This may be explained by one or more possible phenomenon that occur after restricting the analyses to phenotypic extremes, i.e., removing the center of the distribution. First, the extremes of the CPD distribution may be capturing a phenotype more closely approximating physical dependence on nicotine. Tolerance and withdrawal may index severity of nicotine dependence(36), a construct for which we do not have formal diagnoses, but which is highly heritable. In this case, though lacking other important aspects of the clinical presentation such as craving or loss of control, the dichotomized heavy/light phenotype is comparing individuals who may find overnight abstinence less aversive and start smoking later in the day, and endorse lower levels of nicotine dependence (light) to those who meet criteria for severe nicotine dependence (heavy), whereas the standard continuous CPD encoding includes intermediate levels of smoking heaviness that may or may not correlate with clinical presentations of nicotine dependence. Our GREML-based estimate of common, well-tagged *h*^*2*^_*SNP*_ (∼0.09) is approximately the same as one recently reported LDSC-based estimate of nicotine dependence(15), consistent with this hypothesis.

Alternatively, the dichotomized phenotype may reflect lower environmental variance and result in higher *h*^*2*^_*SNP*_, if, for example, environmental effects such as reduced access to cigarettes or regular use of nicotine replacement therapy lead to intermediate values of CPD. Such differences in variance cannot be tested when either trait is dichotomous because the liability underlying the dichotomous trait must be assumed to have unit variance. Ongoing work will seek to distinguish between these two possibilities, and determine whether variants that contribute to heavy/light CPD and other smoking behaviors examined here also contribute to nicotine dependence liability or severity.

We found no evidence of dominance genetic variance for any phenotype, though we note that the power to detect *δ*^*2*^_*SNP*_ is lower relative to *h*^*2*^_*SNP*(19)_ and therefore sample size may be a limiting factor to detect low, but non-zero *δ*^*2*^_*SNP*_. Our findings are consistent with those of Zhu et al.(19), who reported low *δ*^*2*^_*SNP*_ across 79 traits and *δ*^*2*^_*SNP*_∼0 for one smoking phenotype, smoking status. For the four smoking phenotypes in the current study, dominance genetic variance likely contributes little or not at all to the phenotypic variance. We note that dominance interactions of alleles within individual loci may still be contributing to these traits, but as this contributes to additive genetic variance (i.e., *h*^*2*^_*SNP*_), its contribution to *δ*^*2*^_*SNP*_ can be limited, particularly for low-frequency variants(37). Alternatively, interactions between, rather than within, loci may lead to epistatic genetic variation underlying smoking behaviors, and such effects could not be tested using the current approach.

We identified several functional annotations related to LD, MAF, and sequence conservation that significantly contribute to *h*^*2*^_*SNP*_ (Figure 3, Table S5). In addition, GREML-LDMS-I *h*^*2*^_*SNP*_ analyses identified higher contribution of poorly tagged variants relative to well-tagged variants within the same MAF range across all four traits, and also identified nominally significant (95% CI>0) contribution of rare variants (MAF<0.01) for smoking initiation, raw CPD count, and age of initiation. Across the four traits analyzed, rare variants accounted for between 10 and 20% of total *h*^*2*^_*SNP*_ (Table S1). This suggests a role of low-frequency SNPs in low LD with surrounding regions, consistent with purifying and background selection acting to remove mutations with deleterious effects. Given that tobacco use in high concentrations, such as found in cigarettes, is evolutionarily novel for humans, it is unlikely that negative selection acted directly on these smoking behaviors, but rather mutations that today influence nicotine related behaviors may have pleiotropic effects on other traits that were subject to negative selection across evolutionary time(20).

Our *h*^*2*^_*SNP*_ estimates are still considerably lower than twin-based estimates, which range from 50%-80% for dependence, smoking initiation and quantity of use(6-10), suggesting that additional still-missing heritability remains. This is unlikely to be explained by common causal variants, which are well-tagged in current imputation reference panels and from which we expect little downward bias in *h*^*2*^_*SNP*_ estimates(22). Further work will be required to fully characterize non-additive genetic variance, such as epistasis or gene-environment interaction. Rare variants are a likely source of the still-missing heritability. The SE of the rarest MAF partitions were substantially larger than the common variant partition SE, indicating that increased sample size will improve the precision of estimates of rare-variant contribution. Overall, estimates are still generally low compared with those attributable to common variants, and even with large reference panels such as the HRC, rare variants are expected to be poorly imputed, resulting in downwardly-biased *h*^*2*^_*SNP*_(22, 23). Further work through deep sequencing of large samples(38) or using those deeply-sequenced individuals as an improved imputation reference panel is needed to obtain less-biased estimates of rare-variant *h*^*2*^_*SNP*_. For example, height and BMI *h*^*2*^_*SNP*_ estimates using whole genome sequencing have approached twin-based heritability estimates; rare variants account for a substantial proportion of the heritability(39).

Beyond the limitation of rare variant imputation, our study highlights several key issues in *h*^*2*^_*SNP*_ estimation. First, while we used the largest relatively homogenous sample available, even larger samples will be needed for more precise estimation of rare variant contribution, as demonstrated by the much smaller SE of *h*^*2*^_*SNP*_ estimates of traits with larger sample sizes. Second, estimates are sensitive to the estimation method, i.e., H-E Regression-based vs. GREML, which may be due to how environmental confounding differentially influences estimates across methods. GREML-based estimates were relatively stable across relatedness thresholds (Table S1). However, PCGC-based estimates were quite sensitive to relatedness thresholds, being much higher than GREML-based estimates at a .05 threshold and declining with lower thresholds. Although a full assessment of performance of estimators is beyond the scope of this study, it will be important to assess the potential for environmental confounding. As with the possibility of rare variant-environment confounding in GWAS(40), environmental confounding is particularly relevant to estimates of rare variant *h*^*2*^_*SNP*_ because very rare variants are more likely to be shared by individuals sharing recent common ancestors and who may therefore be more likely to share environmental influences. Models that incorporate environmental sharing of families, partners, and close relatives or geography (e.g.,(41, 42)) are a possible avenue to address confounding. To this effect, we note that a full extended twin family design found a lower and possibly sex-dependent estimate of common additive genetic variance, as well as strong environmental influences(18).

In conclusion, though our *h*^*2*^_*SNP*_ estimates of the four different smoking behaviors were generally modest, they are higher than previously published estimates for smoking initiation and cessation and indicate that additional genetic variance may be explained by low- and rare-frequency variants, which may be due to the impact of purifying selection on genes involved in these highly polygenic traits. Quantity of use, as measured by CPD, may also be modestly heritable, but as it depends on the encoding of the variable, additional characterization of the phenotype and its relationship with nicotine dependence is required. All estimates will be improved by the use of complete whole genome sequencing of large numbers of individuals(38), including the contribution of rare variants to smoking behaviors.

## Data Availability

All analyses used data from the UK Biobank.

https://www.ukbiobank.ac.uk/

## Acknowledgements

This work was supported by R01 MH100141-06(PI: Keller); R01 DA 044283, R01 DA 037904, and R01 HG 008983(PI: Vrieze); and the Institute for Behavioral Genetics. We thank John Hewitt, Jerry Stitzel, Charles Hoeffer, Laura Saba, Christian Hopfer, Dana Hancock, Naomi Wray, and Peter Visscher for helpful discussion and comments.

## References

1. US Department of Health and Human Services. Health Consequences of Smoking—50 Years of Progress A Report of the Surgeon General, Report of the Surgeon general 2014: 1081.

2. CENTERS FOR DISEASE CONTROL AND PREVENTION. Quitting Smoking Among Adults — United States, 2001– 2010., Morbidity and Mortality Weekly 2011: 60: 1513–1519.

3. Van Meijgaard J., Fielding J. E. Estimating Benefits of Past, Current, and Future Reductions in Smoking Rates Using a Comprehensive Model With Competing Causes of Death, Preventing Chronic Disease 2012: 110295.

4. Cullen K. A., Ambrose B. K., Gentzke A. S., Apelberg B. J., Jamal A., King B. A. Notes from the Field: Use of Electronic Cigarettes and Any Tobacco Product Among Middle and High School Students - United States, 2011-2018, MMWR Morb Mortal Wkly Rep 2018: 67: 1276–1277.

5. Timpson N. J., Greenwood C. M. T., Soranzo N., Lawson D. J., Richards J. B. Genetic architecture: the shape of the genetic contribution to human traits and disease, Nature Reviews Genetics 2017: 19: 110–124.

6. Haberstick B. C., Ehringer M. A., Lessem J. M., Hopfer C. J., Hewitt J. K. Dizziness and the genetic influences on subjective experiences to initial cigarette use, Addiction 2011: 106: 391–399.

7. Haberstick B. C., Zeiger J. S., Corley R. P., Hopfer C. J., Stallings M. C., Rhee S. H. et al. Common and drug-specific genetic influences on subjective effects to alcohol, tobacco and marijuana use, Addiction 2011: 106: 215–224.

8. Kaprio J. Genetic epidemiology of smoking behavior and nicotine dependence, COPD 2009: 6: 304–306.

9. Rose R.J., Broms U., Korhonen T., Dick D.M., J. K. Genetics of Smoking Behavior. In: Yk K., editor. Handbook of Behavior Genetics, New York, NY: Springer; 2009.

10. Kendler K. S., Schmitt E., Aggen S. H., Prescott C. A., Virginia V. Genetic and Environmental Influences on Alcohol, Caffeine, Cannabis, and Nicotine Use From Early Adolscence to Middle Adulthood., Arch Gen Psychiatry 2008: 65: 674–682.

11. Liu M., Jiang Y., Wedow R., Li Y., Brazel D. M., Chen F. et al. Association studies of up to 1.2 million individuals yield new insights into the genetic etiology of tobacco and alcohol use, Nat Genet 2019: 51: 237–244.

12. TOBACCO and GENETICS CONSORTIUM. Genome-wide meta-analyses identify multiple loci associated with smoking behavior, Nat Genet 2010: 42: 441–447.

13. Hancock D. B., Guo Y., Reginsson G. W., Gaddis N. C., Lutz S. M., Sherva R. et al. Genome-wide association study across European and African American ancestries identifies a SNP in DNMT3B contributing to nicotine dependence, Mol Psychiatry 2018: 23: 1911–1919.

14. Hancock D. B., Wang J. C., Gaddis N. C., Levy J. L., Saccone N. L., Stitzel J. A. et al. A multiancestry study identifies novel genetic associations with CHRNA5 methylation in human brain and risk of nicotine dependence, Hum Mol Genet 2015: 24: 5940–5954.

15. Quach B. C., Bray M. J., Gaddis N. C., Liu M., Palviainen T., Minica C. C. et al. Expanding the genetic architecture of nicotine dependence and its shared genetics with multiple traits: findings from the Nicotine Dependence GenOmics (iNDiGO) Consortium, bioRxiv 2020: DOI:10.1101/2020.1101.1115.898858.

16. Bulik-Sullivan B. K., Loh P. R., Finucane H. K., Ripke S., Yang J., SCHIZOPHRENIA WORKING GROUP OF THE PSYCHIATRIC GENOMICS C. et al. LD Score regression distinguishes confounding from polygenicity in genome-wide association studies, Nat Genet 2015: 47: 291–295.

17. Brazel D. M., Jiang Y., Hughey J. M., Turcot V., Zhan X., Gong J. et al. Exome Chip Meta-analysis Fine Maps Causal Variants and Elucidates the Genetic Architecture of Rare Coding Variants in Smoking and Alcohol Use, Biol Psychiatry 2019: 85: 946–955.

18. Maes H. H., Morley K., Neale M. C., Kendler K. S., Heath A. C., Eaves L. J. et al. Cross-Cultural Comparison of Genetic and Cultural Transmission of Smoking Initiation Using an Extended Twin Kinship Model, Twin Res Hum Genet 2018: 21: 179–190.

19. Zhu Z., Bakshi A., Vinkhuyzen A. A., Hemani G., Lee S. H., Nolte I. M. et al. Dominance genetic variation contributes little to the missing heritability for human complex traits, Am J Hum Genet 2015: 96: 377–385.

20. Schoech A. P., Jordan D. M., Loh P. R., Gazal S., O’Connor L. J., Balick D. J. et al. Quantification of frequency-dependent genetic architectures in 25 UK Biobank traits reveals action of negative selection, Nat Commun 2019: 10: 790.

21. Gazal S., Finucane H. K., Furlotte N. A., Loh P. R., Palamara P. F., Liu X. et al. Linkage disequilibrium-dependent architecture of human complex traits shows action of negative selection, Nat Genet 2017: 49: 1421–1427.

22. Evans L. M., Tahmasbi R., Vrieze S. I., Abecasis G. R., Das S., Gazal S. et al. Comparison of methods that use whole genome data to estimate the heritability and genetic architecture of complex traits, Nature Genetics 2018: 50: 737–745.

23. Yang J., Bakshi A., Zhu Z., Hemani G., Vinkhuyzen A. A. E., Lee S. H. et al. Genetic variance estimation with imputed variants finds negligible missing heritability for human height and body mass index, Nature Genetics 2015: 47: 1114–1120.

24. Evans L. M., Keller M. C. Using partitioned heritability methods to explore genetic architecture, Nature Reviews Genetics 2018: 19: 185–185.

25. Finucane H. K., Bulik-Sullivan B., Gusev A., Trynka G., Reshef Y., Loh P. R. et al. Partitioning heritability by functional annotation using genome-wide association summary statistics, Nat Genet 2015: 47: 1228–1235.

26. Finucane H. K., Reshef Y. A., Anttila V., Slowikowski K., Gusev A., Byrnes A. et al. Heritability enrichment of specifically expressed genes identifies disease-relevant tissues and cell types, Nat Genet 2018: 50: 621–629.

27. Bycroft C., Freeman C., Petkova D., Band G., Elliott L. T., Sharp K. et al. The UK Biobank resource with deep phenotyping and genomic data, Nature 2018: 562: 203–209.

28. Adjangba C., Border R., Romero Villela P. N., Ehringer M. A., Evans L. M. Little Evidence of Modified Genetic Effect of rs16969968 on Heavy Smoking Based on Age of Onset of Smoking, medRxiv 2020: DOI:10.1101/2020.1104.1122.20071407.

29. Heatherton T. F., Kozlowski L. T., Frecker R. C., Fagerstrom K. O. The Fagerstrom Test for Nicotine Dependence: a revision of the Fagerstrom Tolerance Questionnaire, Br J Addict 1991: 86: 1119–1127.

30. Chang C. C., Chow C. C., Tellier L. C., Vattikuti S., Purcell S. M., Lee J. J. Second-generation PLINK: rising to the challenge of larger and richer datasets, Gigascience 2015: 4: 7.

31. Abraham G., Inouye M. Fast principal component analysis of large-scale genome-wide data, PLoS One 2014: 9: e93766.

32. Yang J., Lee S. H., Goddard M. E., Visscher P. M. GCTA: a tool for genome-wide complex trait analysis, Am J Hum Genet 2011: 88: 76–82.

33. Golan D., Lander E. S., Rosset S. Measuring missing heritability: inferring the contribution of common variants, Proc Natl Acad Sci U S A 2014: 111: E5272–5281.

34. Weissbrod O., Flint J., Rosset S. Estimating SNP-Based Heritability and Genetic Correlation in Case-Control Studies Directly and with Summary Statistics, Am J Hum Genet 2018: 103: 89–99.

35. Lee S. H., Yang J., Chen G. B., Ripke S., Stahl E. A., Hultman C. M. et al. Estimation of SNP heritability from dense genotype data, Am J Hum Genet 2013: 93: 1151–1155.

36. Haberstick B. C., Timberlake D., Ehringer M. A., Lessem J. M., Hopfer C. J., Smolen A. et al. Genes, time to first cigarette and nicotine dependence in a general population sample of young adults, Addiction 2007: 102: 655–665.

37. Falconer D. S., Mackay T. F. C. Introduction to quantitative genetics Essex, England: Longman; 1996.

38. Taliun D., Harris D. N., Kessler M. D., Carlson J., Szpiech Z. A., Torres R. et al. Sequencing of 53,831 diverse genomes from the NHLBI TOPMed Program, bioRxiv 2019: DOI:10.1101/563866.

39. Wainschtein P., Jain D. P., Yengo L., Zheng Z., Cupples L. A., Shadyab A. H. et al. Recovery of trait heritability from whole genome sequence data, bioRxiv 2019: DOI:10.1101/588020.

40. Mathieson I., Mcvean G. Differential confounding of rare and common variants in spatially structured populations, Nat Genet 2012: 44: 243–246.

41. Xia C., Amador C., Huffman J., Trochet H., Campbell A., Porteous D. et al. Pedigree- and SNP-Associated Genetics and Recent Environment are the Major Contributors to Anthropometric and Cardiometabolic Trait Variation, PLoS Genet 2016: 12: e1005804.

42. Heckerman D., Gurdasani D., Kadie C., Pomilla C., Carstensen T., Martin H. et al. Linear mixed model for heritability estimation that explicitly addresses environmental variation, Proc Natl Acad Sci U S A 2016: 113: 7377–7382.

